# Magnitude and antibiogram of uropathogens and associated risk factors among asymptomatic female college students in Dessie town, Northeast Ethiopia

**DOI:** 10.1101/2022.09.29.22280506

**Authors:** Berhanu Kebede Reda, Genet Molla, Alemu Gedefie, Daniel Gebretsadik, Mihret Tilahun, Melaku Ashagrie Belete, Agumas Shibabaw

## Abstract

**Background:** Asymptomatic Urinary Tract Infection (Asymptomatic bacteriuria and asymptomatic candiduria) in sexually active non-pregnant female’s population in most cases may not be routinely detected at the initial and reversible stages. This is due to the fact that most women may not feel compelled to seek medical help.

**Objectives:** to determine the prevalence, and factors associated with asymptomatic urinary tract infection, and antibiogram of the uropathogen isolates among asymptomatic female college students

**Methods:** An institutional based cross-sectional study was conducted at selected colleges found in Dessie from January 2021 to March 2021. A total of 422 reproductive age non-pregnant female students were included and socio-demographic and clinical characteristics data were collected using structured questionnaires. Ten ml freshly voided mid-stream urine specimen was collected, transported and processed according to standard operating procedure. Data were coded and entered for statistical analysis using SPSS version 22.0 and descriptive statistics, bivariate and multivariate logistic regression analysis were performed. P-value ≤0.05 with corresponding 95% confidence interval (CI) were considered statistically significant.

**Result:** The overall prevalence of UTI was 24.6%. The prevalence of asymptomatic UTI bacteriuria and candidiuria were 57 (13.5%) and 47 (11.1%) respectively. The predominant isolated uropathogens were *S. saprophyticus* 24 (23.07%) followed by *Candida tropicalis* 23 (22.1%), *Candida albican* 10 (9.61%), *Candida krusei* 9(8.65%) and *E. coli* 8 (7.69%). Gram negative bacterial isolates showed higher level of resistance to Amoxicillin-clavulanic acid 24 (92.3%). Gram positive bacterial uropathogens showed high level of resistance for penicillin 28 (96.6%) and Trimethoprim-Sulfamethoxazole 23 (79.3%). Gram positive bacterial isolates were sensitive to norfloxacin clindamycin and ciprofloxacin accounting 24 (82.7%), 20 (69.0%) and 19 (65.5%), respectively. Moreover, MDR was seen in 50 (87.7%) of all isolated bacterial uropathogens.

Frequency of sexual intercourse (≥ 3 per week) (AOR =7.907, 95% CI: (2.918, 21.425), (P=0.000) and genital area washing habit (during defecation) (AOR = 5.914, 95%CI: (1.860, 18.809), (P=0.0003) and every morning (AOR = 6.128, 95%CI: (1.602, 23.449), (P=0.0008) were found to have statistically significant association with the occurrence of UTI among asymptomatic female college students.

**Conclusion:** A significant prevalence of uropathogens, and high resistance of bacterial isolates to commonly prescribed drugs were observed. Therefore, routine UTI screening, regular health education on the risk of asymptomatic infectious diseases and antimicrobial susceptibility testing should be practiced to avoid the progression of asymptomatic infection into symptomatic UTI.

## INTRODUCTION

Urinary Tract Infection (UTI) is an infection caused by microorganisms at least at one part of the urinary system from the urethra, bladder, ureters and kidneys (1). Asymptomatic uropathogen occurs frequently and is a major cause of UTI (2). It affects millions of people globally, especially in developed countries(3). On a global scale, an estimated 150 million people are affected by bacteriuria each year, either symptomatically or asymptomatically (4)

The organisms causing UTI in a community are most commonly bacteria including *Escherichia coli* which is the commonest, *Proteus mirabilis, Klebsiella* species, *Pseudomonas aeruginosa, Enterobacte*r species, *Enterococci, Citrobacter, Staphylococcus aureus, Staphylococcus saprophyticus, Streptococci* species and other fungi such as *candida* species (5). Women are three times more likely affected by UTI than men, because of shorter urethra, nature of sexual activity, pregnancy, easy contamination of the urinary tract with faecal flora and hormonal changes that occur quickly (6)..

Asymptomatic infection is one which doesn’t show signs of infection while in symptomatic, signs are the major one. Asymptomatic UTI, also known as asymptomatic bacteriuria (ASB) or asymptomatic urinary candidiuria is defined as the presence of significant bacteria (≥ 10^5^ cfu/ml) and a count of ≥ 10^5^ CFU/ml Candida species in an individual ‘s urine without signs and symptoms of UTI respectively (7, 8) Age, sex, sexual activity, and the existence of genitourinary anomalies all play a role in the occurrence of ASB. (9).

Candida species have been identified as one of the common causal agents of UTI caused by fungi. Immunosuppression, extremes of age, diabetes mellitus, structural abnormalities of the urinary tract, broad-spectrum antibiotic use, admission to an intensive care unit (ICU), females in the reproductive age group, indwelling catheters, and abdominal surgeries are the most common risk factors for candiduria. (10). Record affected individuals with candiduria are asymptomatic, and yeasts are noted in the urine as a phenomenon finding on the process of urine culture(11).

Asymptomatic uropathogens causing urinary tract infection among sexually active non-pregnant female population in most cases may not be detected at an initial and reversible stages (12). This is due to the fact that most women may not feel compelled to seek medical help. Significant asymptomatic bacteriuria has been proposed to indicate an early clinical spectrum of urinary tract infection, which could lead to acute infection, chronic infections, or even death due to kidney failure. (13). Even if the infection appears to be minor at first, the asymptomatic patient may develop serious symptoms and pose damage to the part of the urinary tract as the infection advances (14).

Uropathogenic infection of the urinary tract is one of the common reasons for seeking medical attention in the community. Rapidly increasing antimicrobial resistance of uropathogen resulting in limited treatment options. Therefore, knowledge of the uropathogens and their antibiotic susceptibility is important for better treatment of urinary tract infection (15).

Majority of the studies in Africa, particularly Ethiopia, focused on pregnant women and other risk groups including diabetic patients, HIV patients, and pediatric patients, but there is no documented information available on the reproductive age non pregnant women population despite this age group has a higher risk of UTI. Therefore, this study is designed to fill this gap by determining the uropathogens profile and their antibiogram causing asymptomatic UTI among female college students in Dessie town, Northeast Ethiopia.

## MATERIAL AND METHODS

### Study design, area and period

An institution based cross sectional study was conducted from January 2021 - March 2021 in selected colleges in Dessie town, Northeastern Ethiopia. A simple random sampling technique were used to select the colleges included in the study. The study was conducted in Dessie Health Science College (DHSC), Memhran College (MC), Tropical College of Medicine (TCOM) and Alkan Business and Health Science (ABHSC). Dessie Health Science College and MC are governmental colleges whereas TCOM and ABHSC are private colleges. A total of 422 non-pregnant asymptomatic female college students were included using a systematic random sampling technique after proportional allocation was used to determine the number of study participants from the four colleges based on the population size.

### Data and specimen collection

Structured questionnaire was used to obtain information related to demographic characteristic, clinical and risk factors data. About 10ml freshly voided clean-catch midstream urine specimen was collected using pre-labeled (identification code, time, date), wide mouth, leak-proof, sterile, screw-capped plastic container (FL Medical, Italy) by study participants after appropriate specimen collection instructions were given.

### Specimen transportation

The collected specimens were immediately transported to Wollo University Microbiology Laboratory in a cold box for processing, after confirming their non-pregnancy by using human chorionic gonadotropin (HCG) pregnancy testing kit. In case of unavoidable delay, very few specimens were kept refrigerated at 4 °C until being processed. Immediate inoculation had been performed for the rest of the specimens on arrival to the laboratory.

### Cultivation and identification of isolates

Using calibrated wire loop (0.001ml) urine specimen was inoculated in to Cystine Lactose Electrolyte Deficient medium (CLED) (Oxoid Ltd, UK). Inoculated culture media were then incubated overnight in aerobic atmosphere at 37°C for 24 hours. Colonies were counted to check the presence of significant growth. Colony counts yielding bacterial growth of ≥10^5^ CFU/ml of urine was regarded as significant bacteriuria (SB); but specimens that produce <10^5^CFU/ml were considered insignificant or due to contamination (32).

Based on their gram staining reaction, colonies from CLED agar were sub cultured into MacConkey agar (Oxoid, Ltd), blood agar plates and Mannitol salt agar (Hi Media™), then incubated at 37°C for 24 hours. Identification of bacterial species was done using colony characteristics, gram staining reaction and biochemical tests following standard procedure. The gram-negative bacteria were identified by indole production, motility test, citrate utilization, urease test, H_2_S production in Kligler Iron Agar (KIA) and carbohydrate utilization tests. The gram-positive bacteria was identified using catalase, coagulase tests and novobiocin susceptibility test (32).

For fungal identification a loopful (0.001 ml) of well-mixed un-centrifuged urine was streak onto the surface of Sabouraud dextrose agar (SDA) (32). The plates were incubated aerobically at 37◦C for 24-48 hours and counts were expressed in CFU/ml. A count of ≥ 10^5^ CFU/ml was considered indicative of asymptomatic urinary candidiuria (8). The significant fungal isolates were sub cultured on to CHROM agar TM Candida media (chrome agar, Paris, France). All fungal isolates were identified with CHROM agar TM Candida Media to confirm the presence of the five *Candida species (C*.*albican, C*.*tropicalis, C*.*krusei, C*.*glabrta and C*.*auris) (33)*. Furthermore, all uropathogen isolates were identified using standard clinical laboratory methods

### Antimicrobial Susceptibility Testing

The antimicrobial susceptibility patterns of all identified bacterial isolates were performed according to the criteria of Clinical and Laboratory Standards Institute (34) using the Kirby-Bauer disc diffusion method on Muller-Hinton Agar. A loop full of bacteria was taken from a pure culture colony and transferred to a tube containing 5ml of normal saline and mixed gently until it forms a homogenous suspension. The turbidity of the suspension was adjusted to the turbidity of 0.5 McFarland in order to standardize the inoculum size. A sterile cotton swab was dipped, rotated across the wall of the tube to avoid excess fluid and was evenly inoculated on Muller-Hinton agar (Conda ltd, USA) and then the antibiotic discs were placed on MHA plates.

The following antimicrobials were used clindamycin (CL, 10μg), penicillin (PEN, 10µg), chloramphenicol, (CAF, 30μg), ciprofloxacin (CIP, 5µg), Tetracycline (TTC, 30µg), trimethoprim sulfamethoxazole (SXT, 1.25/23.75µg), nitrofurantoin (F, 300µg), and norfloxacin (NOR, 10µg) for Gram positives; on the other hand ciprofloxacin (CIP, 5µg), tetracycline (TTC, 30µg), trimethoprim sulfamethoxazole (SXT, 1.25/23.75µg), nitrofurantoin (F, 300µg), norfloxacin (NOR, 10µg), ceftriaxone (CRO, 30μg), amoxicillin-clavulanic acid (AMC, 20/10μg), cefotaxime (CTX, 30µg), ceftazidime (CAZ, 30µg), ampicillin (AMP, 10μg), amikacin (AMK, 30μg) and gentamicin (GN, 10μg) were used for Gram negative bacteria. These antimicrobial drug disks were selected based on Clinical and Laboratory Standards Institute (CLSI) and also by considering the availability and frequent prescriptions of these drugs for the treatment of UTIs in the study area.

The plates were overnight incubated at 37°C. Diameters of the zone of inhibition around the discs was measured using a digital caliper. The interpretation of the results of the antimicrobial susceptibility tests was based on the standardized table supplied by the national committee for CLSI (34) criteria as sensitive, resistant and intermediate as a resistant.

### Quality assurance

All quality control tests were performed before, during, and after data collection to ensure that the data was of high quality and reliability. Every question in the structured questionnaire was written in a clear and explicit manner and then translated into the local language (Amharic). Training was given for data collectors on how to collect data. The completeness of the questionnaires was checked by principal investigator. The questionnaire was pretested on 5% of the sample size (22 asymptomatic female students) from Mankul college which is located in Dessie city, Northeast Ethiopia. Moreover, all laboratory assays were done by maintaining the quality control procedures. The performance of Gram staining chemical was checked by colonies of *S. aureus* (ATCC 25923) and *E. coli* (ATCC 25922). Standard Operating Procedures (SOPs) were strictly followed verifying that media meet expiration date and quality control parameters per CLSI guideline. Biochemical media were evaluated using positive and negative control organisms. Visual inspections of holes, uneven filling, and haemolysis, signs of freezing, bubbles and corrosion in media or plastic Petri dishes was conducted. standardized inoculum density of bacterial suspension was prepared, saline turbidity standard equivalent to a 0.5 McFarland standard was used to control the quality of antimicrobial susceptibility test. Standard reference strain of *S. aureus* (ATCC-25923), *E. coli* (ATCC-25922), *K. pneumoniae* (ATCC 700603) and *C. albican* (ATCC 90028) were used as quality control strains throughout the study. The inhibition diameter zone interpretation was carried out on the basis of the updated 2020 CLSI guideline.

### Statistical analysis

The data generated were entered and analyzed by Statistical Package for Social Sciences (SPSS) version 22. (IBM, USA). Descriptive statistics was computed and data were presented using texts, figures and tables. To show the relationship between each variable and the dependent variable, binary logistic regression was utilized. Moreover, a multivariate analysis was employed to identify factors that independently influence the occurrence of the dependent variable. P-value ≤ 0.05 with 95% confidence interval was considered statistically significant.

### Ethical approval and consent to participate

The ethical approval was obtained from College of Medicine and Health Science Research Ethics Review Committee (RERC), Wollo University. Official cooperation and permission were obtained from selected colleges in Dessie town. Moreover, prior to commencing the study, a written informed consent was obtained from each study participant. Subject confidentiality and any special data security requirements were maintained and assured. Results of the laboratory examinations that have a direct benefit in the health of the study participants were linked to a health facility and the participants get their results and treatment duly as required. Moreover, this study was conducted in accordance with the Declaration of Helsinki.

## RESULTS

### Socio-demographic characteristics

A total of 422 female college students were investigated for asymptomatic UTI. The median age of the study participants was 21 years, ranged from 18 - 38 years. Majority of the study participants 209 (49.4%) were in the age group of 21 to 25 years (Table 1). Student batch of participants varied from second year up to fourth year and around 177 (41.9 %) of study participants were second year students.

**Table 1:**
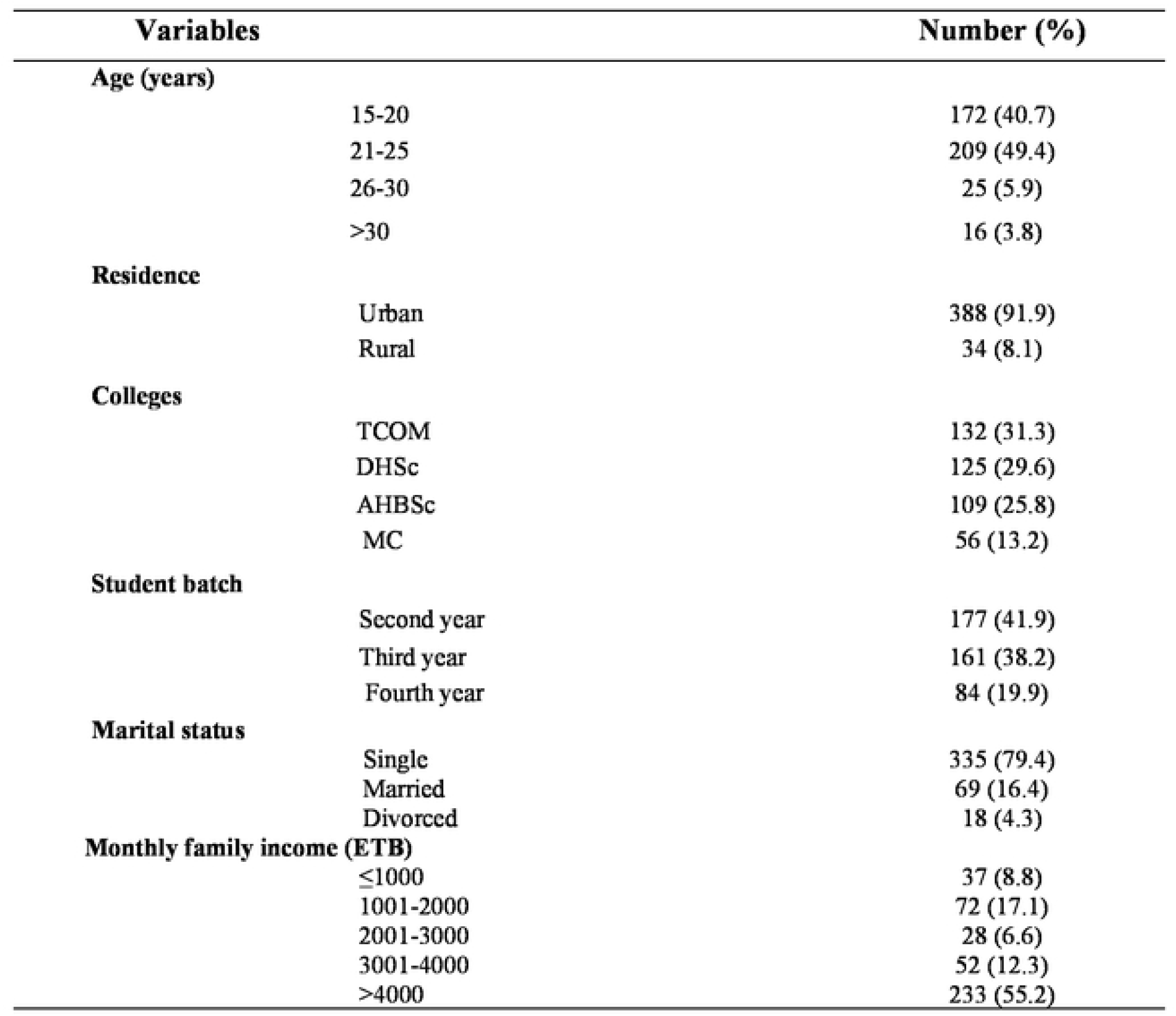
Socio-demographic characteristics of the study participants in Dessie town, Northeast Ethiopia, January 2021 - March 2021.

### Prevalence of urinary tract infection

The prevalence of significant bacteriuria and candiduria was 57 (13.5%) and 47 (11.1%), respectively. The overall prevalence of UTI was 24.6% (104/422) (Figure 1).

**Figure 1:**
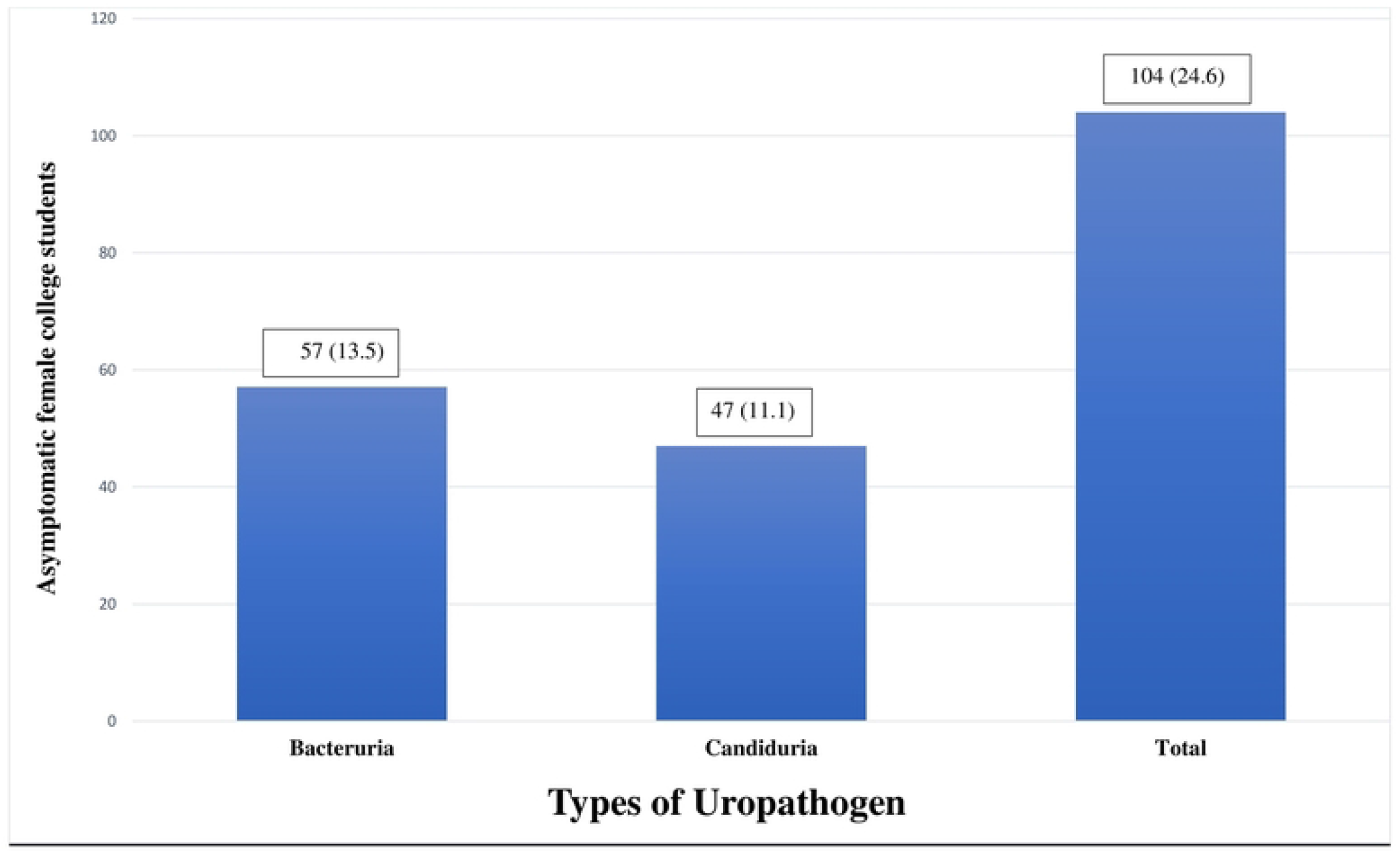
Prevalence of urinary tract infection among asymptomatic female college students in Dessie town, Northeast Ethiopia, January 2021 - March 2021.

### Associated risk factors of UTI

In this study, 15 independent variables were considered during the bivariate analysis of risk factors for bacterial and candidial UTI. In multivariate analysis, frequency of sexual intercourse (AOR =7.907, 95% CI: (2.918, 21.425), P<0.001) and genital area washing habit (During defecation) AOR = 5.914, 95%CI: (1.860, 18.809), P=0.003) and every morning AOR = 6.128, 95%CI: (1.602, 23.449), P=0.008) were found to have statistically significant association with the occurrence of UTI. Out of a total of 94 female students who had significant uropathogen, 13 (13.8%) had >= 3 sexual frequency activity (P<0.001), and 45 (47.9) (P=0.003) and 13 (13.8) (P=0.008) had an after defecation and every morning genital area washing habit respectively (Table 2).

**Table 2:**
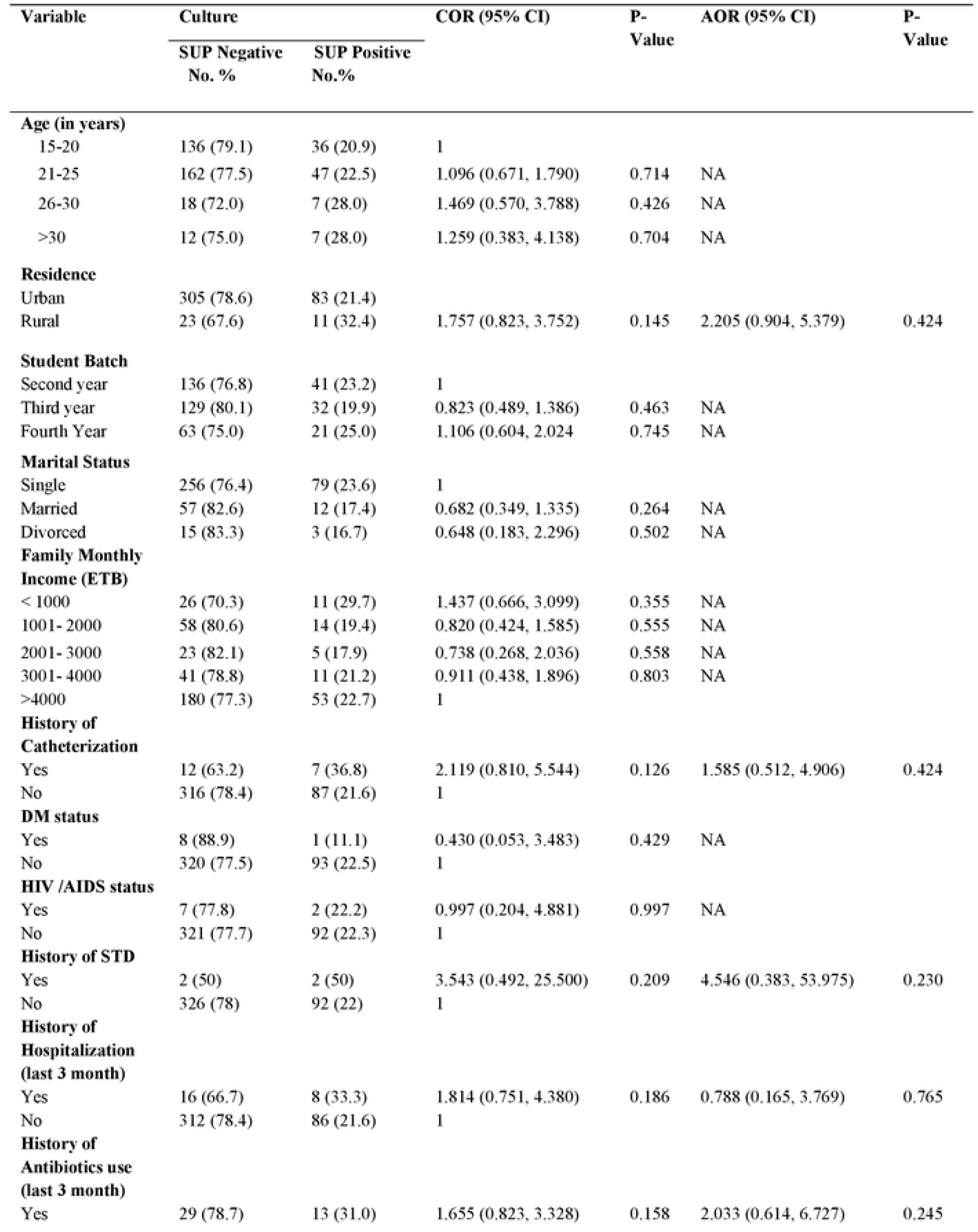
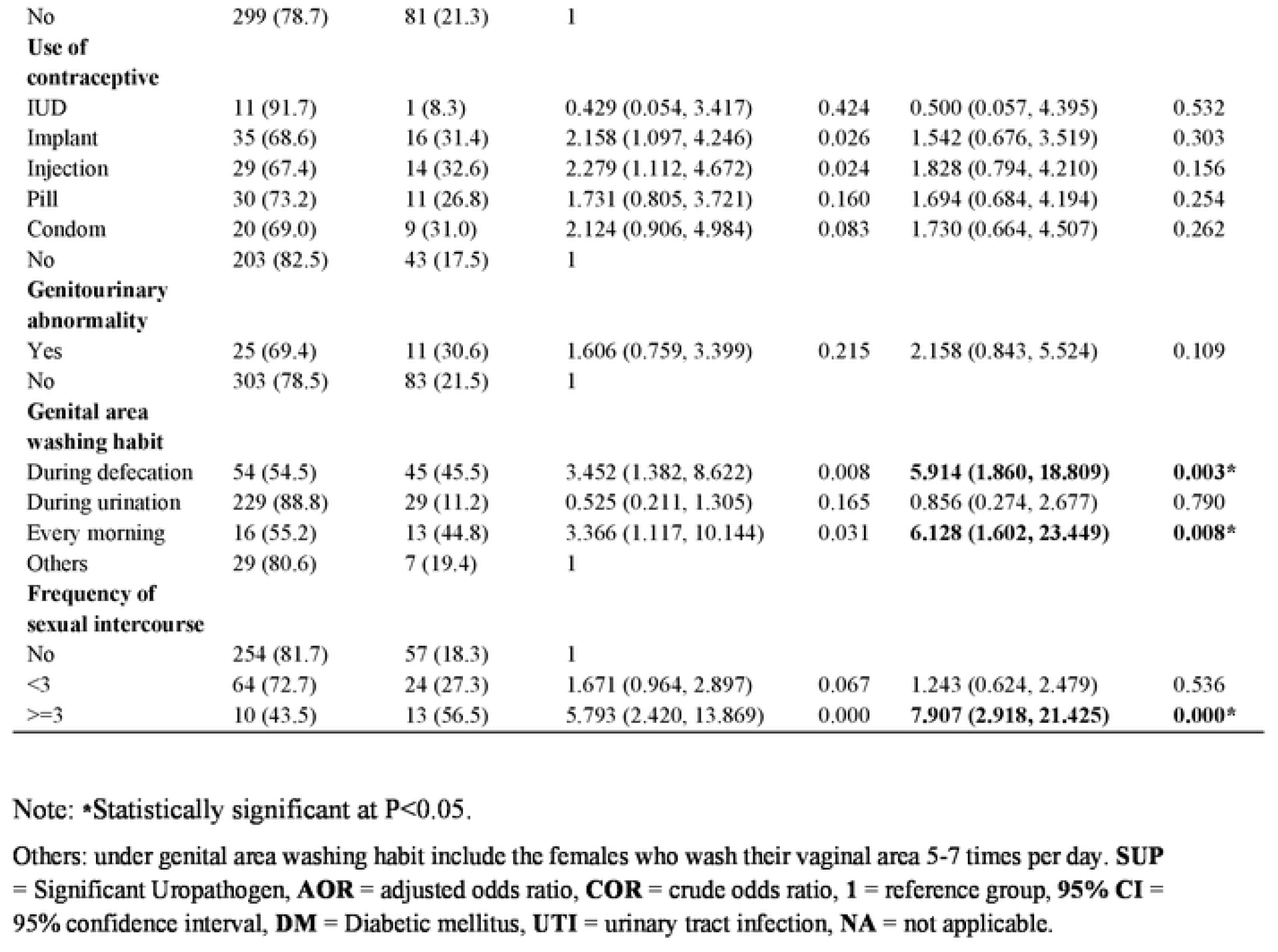
Bivariate and Multivariate logistic regression analysis of risk factors of asymptomatic uropathogens among female college students in Dessie town, Northeast Ethiopia, January 2021 - March 2021.

| Variable | Culture |  | COR (95% CI) | P-Value | AOR (95% CI) | P-Value |
| --- | --- | --- | --- | --- | --- | --- |
|  | SUP Negative<br>No. % | SUP Positive<br>No.% |  |  |  |  |
| **Age (in years)** |  |  |  |  |  |  |
| 15-20 | 136 (79.1) | 36 (20.9) | 1 |  |  |  |
| 21-25 | 162 (77.5) | 47 (22.5) | 1.096 (0.671, 1.790) | 0.714 | NA |  |
| 26-30 | 18 (72.0) | 7 (28.0) | 1.469 (0.570, 3.788) | 0.426 | NA |  |
| >30 | 12 (75.0) | 7 (28.0) | 1.259 (0.383, 4.138) | 0.704 | NA |  |
| **Residence** |  |  |  |  |  |  |
| Urban | 305 (78.6) | 83 (21.4) |  |  |  |  |
| Rural | 23 (67.6) | 11 (32.4) | 1.757 (0.823, 3.752) | 0.145 | 2.205 (0.904, 5.379) | 0.424 |
| **Student Batch** |  |  |  |  |  |  |
| Second year | 136 (76.8) | 41 (23.2) | 1 |  |  |  |
| Third year | 129 (80.1) | 32 (19.9) | 0.823 (0.489, 1.386) | 0.463 | NA |  |
| Fourth Year | 63 (75.0) | 21 (25.0) | 1.106 (0.604, 2.024 | 0.745 | NA |  |
| **Marital Status** |  |  |  |  |  |  |
| Single | 256 (76.4) | 79 (23.6) | 1 |  |  |  |
| Married | 57 (82.6) | 12 (17.4) | 0.682 (0.349, 1.335) | 0.264 | NA |  |
| Divorced | 15 (83.3) | 3 (16.7) | 0.648 (0.183, 2.296) | 0.502 | NA |  |
| **Family Monthly Income (ETB)** |  |  |  |  |  |  |
| < 1000 | 26 (70.3) | 11 (29.7) | 1.437 (0.666, 3.099) | 0.355 | NA |  |
| 1001- 2000 | 58 (80.6) | 14 (19.4) | 0.820 (0.424, 1.585) | 0.555 | NA |  |
| 2001- 3000 | 23 (82.1) | 5 (17.9) | 0.738 (0.268, 2.036) | 0.558 | NA |  |
| 3001- 4000 | 41 (78.8) | 11 (21.2) | 0.911 (0.438, 1.896) | 0.803 | NA |  |
| >4000 | 180 (77.3) | 53 (22.7) | 1 |  |  |  |
| **History of Catheterization** |  |  |  |  |  |  |
| Yes | 12 (63.2) | 7 (36.8) | 2.119 (0.810, 5.544) | 0.126 | 1.585 (0.512, 4.906) | 0.424 |
| No | 316 (78.4) | 87 (21.6) | 1 |  |  |  |
| **DM status** |  |  |  |  |  |  |
| Yes | 8 (88.9) | 1 (11.1) | 0.430 (0.053, 3.483) | 0.429 | NA |  |
| No | 320 (77.5) | 93 (22.5) | 1 |  |  |  |
| **HIV /AIDS status** |  |  |  |  |  |  |
| Yes | 7 (77.8) | 2 (22.2) | 0.997 (0.204, 4.881) | 0.997 | NA |  |
| No | 321 (77.7) | 92 (22.3) | 1 |  |  |  |
| **History of STD** |  |  |  |  |  |  |
| Yes | 2 (50) | 2 (50) | 3.543 (0.492, 25.500) | 0.209 | 4.546 (0.383, 53.975) | 0.230 |
| No | 326 (78) | 92 (22) | 1 |  |  |  |
| **History of Hospitalization (last 3 month)** |  |  |  |  |  |  |
| Yes | 16 (66.7) | 8 (33.3) | 1.814 (0.751, 4.380) | 0.186 | 0.788 (0.165, 3.769) | 0.765 |
| No | 312 (78.4) | 86 (21.6) | 1 |  |  |  |
| **History of Antibiotics use (last 3 month)** |  |  |  |  |  |  |
| Yes | 29 (78.7) | 13 (31.0) | 1.655 (0.823, 3.328) | 0.158 | 2.033 (0.614, 6.727) | 0.245 |
| No | 299 (78.7) | 81 (21.3) | 1 |  |  |  |
| **Use of**<br>**contraceptive** |  |  |  |  |  |  |
| IUD | 11 (91.7) | 1 (8.3) | 0.429 (0.054, 3.417) | 0.424 | 0.500 (0.057, 4.395) | 0.532 |
| Implant | 35 (68.6) | 16 (31.4) | 2.158 (1.097, 4.246) | 0.026 | 1.542 (0.676, 3.519) | 0.303 |
| Injection | 29 (67.4) | 14 (32.6) | 2.279 (1.112, 4.672) | 0.024 | 1.828 (0.794, 4.210) | 0.156 |
| Pill | 30 (73.2) | 11 (26.8) | 1.731 (0.805, 3.721) | 0.160 | 1.694 (0.684, 4.194) | 0.254 |
| Condom | 20 (69.0) | 9 (31.0) | 2.124 (0.906, 4.984) | 0.083 | 1.730 (0.664, 4.507) | 0.262 |
| No | 203 (82.5) | 43 (17.5) | 1 |  |  |  |
| **Genitourinary**<br>**abnormality** |  |  |  |  |  |  |
| Yes | 25 (69.4) | 11 (30.6) | 1.606 (0.759, 3.399) | 0.215 | 2.158 (0.843, 5.524) | 0.109 |
| No | 303 (78.5) | 83 (21.5) | 1 |  |  |  |
| **Genital area**<br>**washing habit** |  |  |  |  |  |  |
| During defecation | 54 (54.5) | 45 (45.5) | 3.452 (1.382, 8.622) | 0.008 | **5.914 (1.860, 18.809)** | **0.003\*** |
| During urination | 229 (88.8) | 29 (11.2) | 0.525 (0.211, 1.305) | 0.165 | 0.856 (0.274, 2.677) | 0.790 |
| Every morning | 16 (55.2) | 13 (44.8) | 3.366 (1.117, 10.144) | 0.031 | **6.128 (1.602, 23.449)** | **0.008\*** |
| Others | 29 (80.6) | 7 (19.4) | 1 |  |  |  |
| **Frequency of**<br>**sexual intercourse** |  |  |  |  |  |  |
| No | 254 (81.7) | 57 (18.3) | 1 |  |  |  |
| <3 | 64 (72.7) | 24 (27.3) | 1.671 (0.964, 2.897) | 0.067 | 1.243 (0.624, 2.479) | 0.536 |
| >=3 | 10 (43.5) | 13 (56.5) | 5.793 (2.420, 13.869) | 0.000 | **7.907 (2.918, 21.425)** | **0.000\*** |

Note: \*Statistically significant at P<0.05.
Others: under genital area washing habit include the females who wash their vaginal area 5-7 times per day. **SUP** = Significant Uropathogen, **AOR** = adjusted odds ratio, **COR** = crude odds ratio, **1** = reference group, **95% CI** = 95% confidence interval, **DM** = Diabetic mellitus, **UTI** = urinary tract infection, **NA** = not applicable.

### Bacterial and fungal uropathogens isolates

A total of 13 bacteria and 4 candida species were isolated from mid-stream urine sample. Almost all 103 (99.0%) of the infected female students had single infection; while only 1 (0.96%) was dually infected (*C. albican and C. krusei*), which makes the total number of bacterial isolates 57 and fungal isolates 47 (Table 3). The predominantly isolated uropathogen was *S. saprophyticus* 24 (23.07%) followed by *Candida tropicalis* 23 (22.1%), *Candida albican* 10 (9.61%), *Candida krusei* 9(8.65%) and *E. coli* 8 (7.69%).

**Table 3:** Frequency of bacterial and fungal uropathogens of asymptomatic UTI among female college students in Dessie town, Northeast Ethiopia, January 2021 - March 2021.

| Uropathogen isolates | N (%) from 104 |
| --- | --- |
| **Bacterial isolates** |  |
| **Gram positive** |  |
| *S. saprophyticus* | 24 (23.07) |
| *S. aureus* | 4 (3.84) |
| *Staphylococcus epidermidis* | 1 (0.96) |
| **Gram negative** |  |
| *E. coli* | 8 (7.69) |
| *Klebsiella ozenae* | 5 (1.92) |
| *Klebsiella rhinoscleromatis* | 3 (2.88) |
| *Providencia stuarti* | 3 (2.88) |
| *Citrobacter* | 2 (3.5) |
| *Serratia* | 2 (1.92) |
| *Citrobacter diversus* | 2 (1.92) |
| *Acinetobacter spp* | 1 (0.96) |
| *Klebsella oxytoca* | 1 (0.96) |
| *Klebsiella pneumonia* | 1 (0.96) |
| **Sub total** | **57 (54.8)** |
| **Fungi isolates** |  |
| *Candida tropicalis* | 23 (22.11) |
| *Candida albican* | 10 (9.61) |
| *Candida krusei* | 9 (8.65) |
| *Candida galberta* | 6 (5.76) |
| **Sub total** | **47 (45.19)** |
| **Total** | **104 (100)** |

### Antimicrobial susceptibility pattern of bacterial uropathogens

Majority of the isolated Gram negative uropathogens showed resistance for amoxicillin-clavulanic acid (92.3%). Rates of resistance of Gram-negatives against tetracycline, trimethoprim-sulfamethoxazole, ampicillin ranged from 52.0% – 71.4%. However, all Gram-negative bacterial isolates showed relatively low level of resistance against norfloxacin 7.1%, gentamicin 8% and ciprofloxacin 10.7% (Table 4).

**Table 2:**
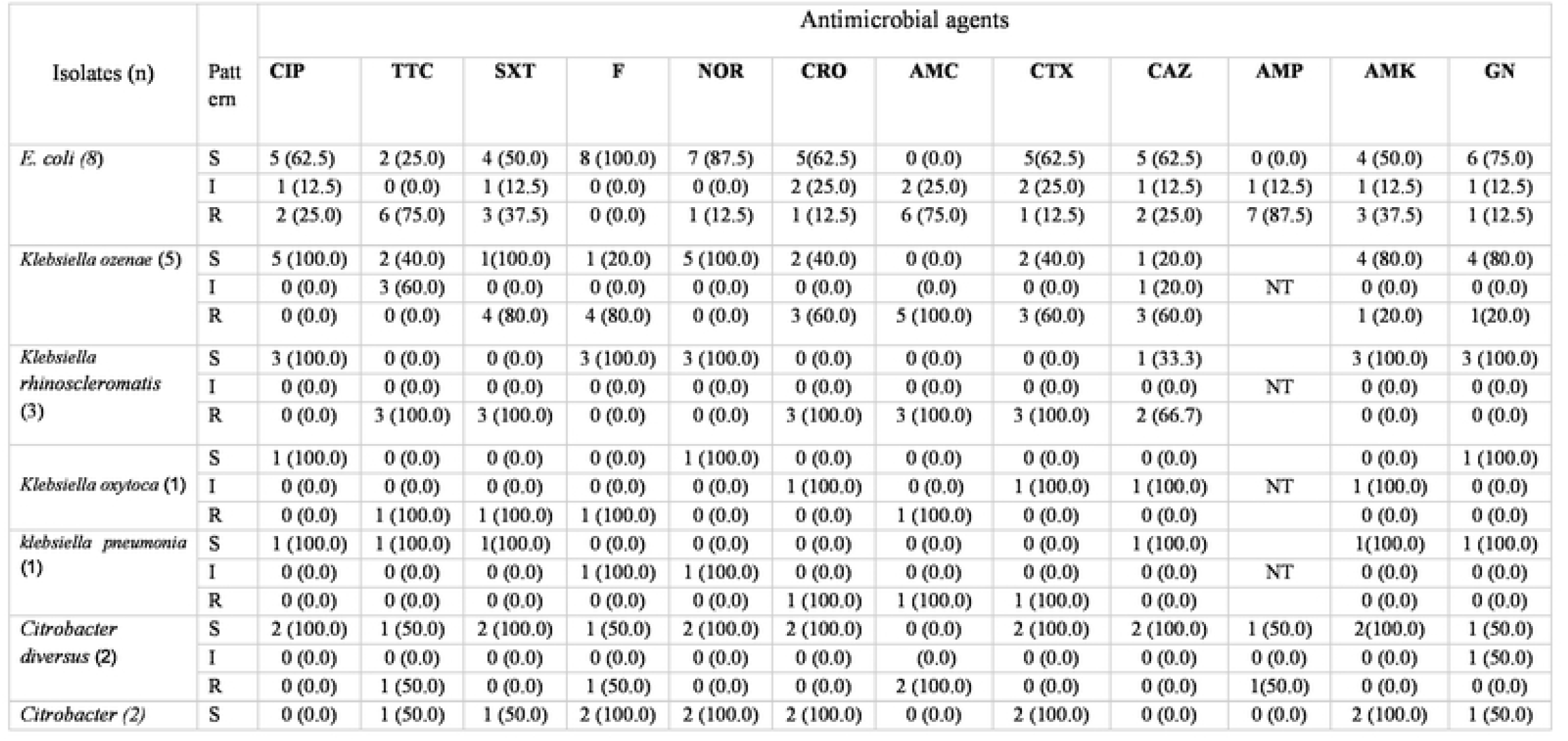
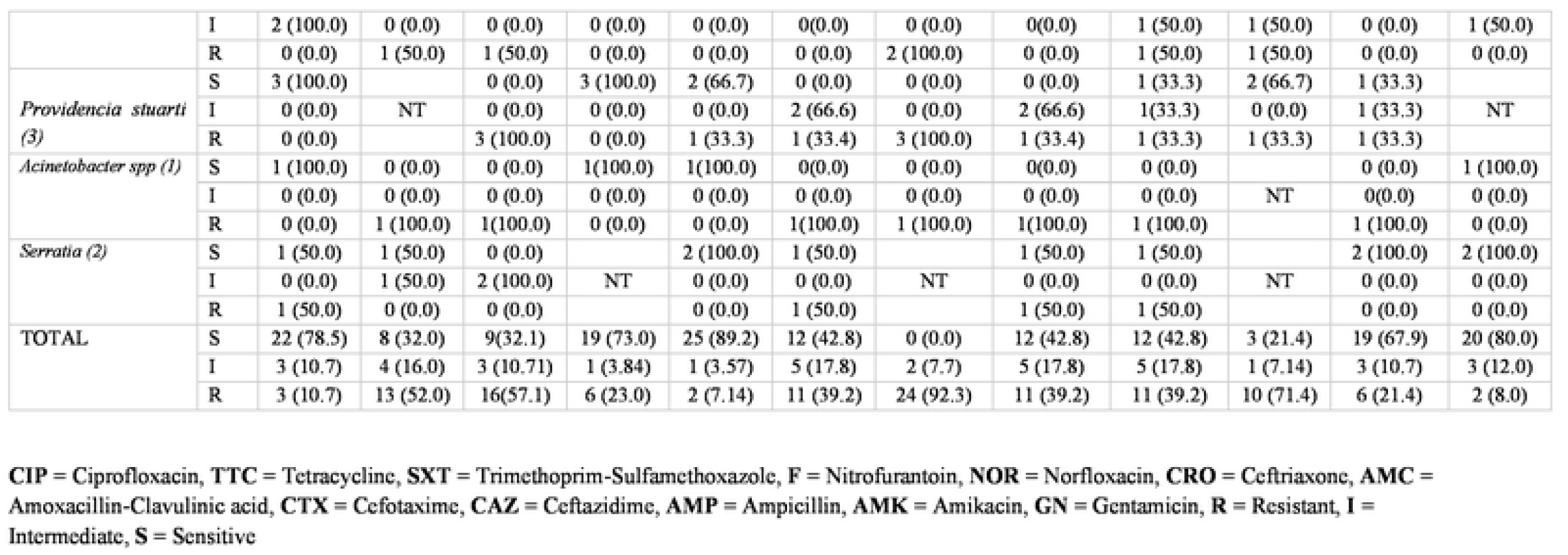
Antimicrobial susceptibility pattern of Gram-negative bacteria (n=28) isolated from urine culture among Asymptomatic female college students at Dessie town, Northeast Ethiopia, January 2021 - March 2021.

| Isolates (n) | Pattern | Antimicrobial agents |  |  |  |  |  |  |  |  |  |  |  |
| --- | --- | --- | --- | --- | --- | --- | --- | --- | --- | --- | --- | --- | --- |
|  |  | CIP | TTC | SXT | F | NOR | CRO | AMC | CTX | CAZ | AMP | AMK | GN |
| *E. coli* (8) | S | 5 (62.5) | 2 (25.0) | 4 (50.0) | 8 (100.0) | 7 (87.5) | 5(62.5) | 0 (0.0) | 5(62.5) | 5 (62.5) | 0 (0.0) | 4 (50.0) | 6 (75.0) |
|  | I | 1 (12.5) | 0 (0.0) | 1 (12.5) | 0 (0.0) | 0 (0.0) | 2 (25.0) | 2 (25.0) | 2 (25.0) | 1 (12.5) | 1 (12.5) | 1 (12.5) | 1 (12.5) |
|  | R | 2 (25.0) | 6 (75.0) | 3 (37.5) | 0 (0.0) | 1 (12.5) | 1 (12.5) | 6 (75.0) | 1 (12.5) | 2 (25.0) | 7 (87.5) | 3 (37.5) | 1 (12.5) |
| *Klebsiella ozenae* (5) | S | 5 (100.0) | 2 (40.0) | 1(100.0) | 1 (20.0) | 5 (100.0) | 2 (40.0) | 0 (0.0) | 2 (40.0) | 1 (20.0) |  | 4 (80.0) | 4 (80.0) |
|  | I | 0 (0.0) | 3 (60.0) | 0 (0.0) | 0 (0.0) | 0 (0.0) | 0 (0.0) | (0.0) | 0 (0.0) | 1 (20.0) | NT | 0 (0.0) | 0 (0.0) |
|  | R | 0 (0.0) | 0 (0.0) | 4 (80.0) | 4 (80.0) | 0 (0.0) | 3 (60.0) | 5 (100.0) | 3 (60.0) | 3 (60.0) |  | 1 (20.0) | 1(20.0) |
| *Klebsiella rhinoscleromatis* (3) | S | 3 (100.0) | 0 (0.0) | 0 (0.0) | 3 (100.0) | 3 (100.0) | 0 (0.0) | 0 (0.0) | 0 (0.0) | 1 (33.3) |  | 3 (100.0) | 3 (100.0) |
|  | I | 0 (0.0) | 0 (0.0) | 0 (0.0) | 0 (0.0) | 0 (0.0) | 0 (0.0) | 0 (0.0) | 0 (0.0) | 0 (0.0) | NT | 0 (0.0) | 0 (0.0) |
|  | R | 0 (0.0) | 3 (100.0) | 3 (100.0) | 0 (0.0) | 0 (0.0) | 3 (100.0) | 3 (100.0) | 3 (100.0) | 2 (66.7) |  | 0 (0.0) | 0 (0.0) |
| *Klebsiella oxytoca* (1) | S | 1 (100.0) | 0 (0.0) | 0 (0.0) | 0 (0.0) | 1 (100.0) | 0 (0.0) | 0 (0.0) | 0 (0.0) | 0 (0.0) |  | 0 (0.0) | 1 (100.0) |
|  | I | 0 (0.0) | 0 (0.0) | 0 (0.0) | 0 (0.0) | 0 (0.0) | 1 (100.0) | 0 (0.0) | 1 (100.0) | 1 (100.0) | NT | 1 (100.0) | 0 (0.0) |
|  | R | 0 (0.0) | 1 (100.0) | 1 (100.0) | 1 (100.0) | 0 (0.0) | 0 (0.0) | 1 (100.0) | 0 (0.0) | 0 (0.0) |  | 0 (0.0) | 0 (0.0) |
| *klebsiella pneumonia* (1) | S | 1 (100.0) | 1 (100.0) | 1(100.0) | 0 (0.0) | 0 (0.0) | 0 (0.0) | 0 (0.0) | 0 (0.0) | 1 (100.0) |  | 1(100.0) | 1 (100.0) |
|  | I | 0 (0.0) | 0 (0.0) | 0 (0.0) | 1 (100.0) | 1 (100.0) | 0 (0.0) | 0 (0.0) | 0 (0.0) | 0 (0.0) | NT | 0 (0.0) | 0 (0.0) |
|  | R | 0 (0.0) | 0 (0.0) | 0 (0.0) | 0 (0.0) | 0 (0.0) | 1 (100.0) | 1 (100.0) | 1 (100.0) | 0 (0.0) |  | 0 (0.0) | 0 (0.0) |
| *Citrobacter diversus* (2) | S | 2 (100.0) | 1 (50.0) | 2 (100.0) | 1 (50.0) | 2 (100.0) | 2 (100.0) | 0 (0.0) | 2 (100.0) | 2 (100.0) | 1 (50.0) | 2(100.0) | 1 (50.0) |
|  | I | 0 (0.0) | 0 (0.0) | 0 (0.0) | 0 (0.0) | 0 (0.0) | 0 (0.0) | (0.0) | 0 (0.0) | 0 (0.0) | 0 (0.0) | 0 (0.0) | 1 (50.0) |
|  | R | 0 (0.0) | 1 (50.0) | 0 (0.0) | 1 (50.0) | 0 (0.0) | 0 (0.0) | 2 (100.0) | 0 (0.0) | 0 (0.0) | 1(50.0) | 0 (0.0) | 0 (0.0) |
| *Citrobacter* (2) | S | 0 (0.0) | 1 (50.0) | 1 (50.0) | 2 (100.0) | 2 (100.0) | 2 (100.0) | 0 (0.0) | 2 (100.0) | 0 (0.0) | 0 (0.0) | 2 (100.0) | 1 (50.0) |
|  | I | 2 (100.0) | 0 (0.0) | 0 (0.0) | 0 (0.0) | 0 (0.0) | 0(0.0) | 0 (0.0) | 0(0.0) | 1 (50.0) | 1 (50.0) | 0 (0.0) | 1 (50.0) |
|  | R | 0 (0.0) | 1 (50.0) | 1 (50.0) | 0 (0.0) | 0 (0.0) | 0 (0.0) | 2 (100.0) | 0 (0.0) | 1 (50.0) | 1 (50.0) | 0 (0.0) | 0 (0.0) |
| *Providencia stuarti*<br>(3) | S | 3 (100.0) |  | 0 (0.0) | 3 (100.0) | 2 (66.7) | 0 (0.0) | 0 (0.0) | 0 (0.0) | 1 (33.3) | 2 (66.7) | 1 (33.3) |  |
|  | I | 0 (0.0) | NT | 0 (0.0) | 0 (0.0) | 0 (0.0) | 2 (66.6) | 0 (0.0) | 2 (66.6) | 1(33.3) | 0 (0.0) | 1 (33.3) | NT |
|  | R | 0 (0.0) |  | 3 (100.0) | 0 (0.0) | 1 (33.3) | 1 (33.4) | 3 (100.0) | 1 (33.4) | 1 (33.3) | 1 (33.3) | 1 (33.3) |  |
| *Acinetobacter spp* (1) | S | 1 (100.0) | 0 (0.0) | 0 (0.0) | 1(100.0) | 1(100.0) | 0(0.0) | 0 (0.0) | 0(0.0) | 0 (0.0) |  | 0 (0.0) | 1 (100.0) |
|  | I | 0 (0.0) | 0 (0.0) | 0 (0.0) | 0 (0.0) | 0 (0.0) | 0 (0.0) | 0 (0.0) | 0 (0.0) | 0 (0.0) | NT | 0(0.0) | 0 (0.0) |
|  | R | 0 (0.0) | 1 (100.0) | 1(100.0) | 0 (0.0) | 0 (0.0) | 1(100.0) | 1 (100.0) | 1(100.0) | 1 (100.0) |  | 1 (100.0) | 0 (0.0) |
| *Serratia* (2) | S | 1 (50.0) | 1 (50.0) | 0 (0.0) |  | 2 (100.0) | 1 (50.0) |  | 1 (50.0) | 1 (50.0) |  | 2 (100.0) | 2 (100.0) |
|  | I | 0 (0.0) | 1 (50.0) | 2 (100.0) | NT | 0 (0.0) | 0 (0.0) | NT | 0 (0.0) | 0 (0.0) | NT | 0 (0.0) | 0 (0.0) |
|  | R | 1 (50.0) | 0 (0.0) | 0 (0.0) |  | 0 (0.0) | 1 (50.0) |  | 1 (50.0) | 1 (50.0) |  | 0 (0.0) | 0 (0.0) |
| TOTAL | S | 22 (78.5) | 8 (32.0) | 9(32.1) | 19 (73.0) | 25 (89.2) | 12 (42.8) | 0 (0.0) | 12 (42.8) | 12 (42.8) | 3 (21.4) | 19 (67.9) | 20 (80.0) |
|  | I | 3 (10.7) | 4 (16.0) | 3 (10.71) | 1 (3.84) | 1 (3.57) | 5 (17.8) | 2 (7.7) | 5 (17.8) | 5 (17.8) | 1 (7.14) | 3 (10.7) | 3 (12.0) |
|  | R | 3 (10.7) | 13 (52.0) | 16(57.1) | 6 (23.0) | 2 (7.14) | 11 (39.2) | 24 (92.3) | 11 (39.2) | 11 (39.2) | 10 (71.4) | 6 (21.4) | 2 (8.0) |

CIP = Ciprofloxacin, TTC = Tetracycline, SXT = Trimethoprim-Sulfamethoxazole, F = Nitrofurantoin, NOR = Norfloxacin, CRO = Ceftriaxone, AMC = Amoxicillin-Clavulinic acid, CTX = Cefotaxime, CAZ = Ceftazidime, AMP = Ampicillin, AMK = Amikacin, GN = Gentamicin, R = Resistant, I = Intermediate, S = Sensitive

In general, Gram-positive bacterial isolates showed high level of resistance for penicillin 28 (96.6%) and trimethoprim-sulfamethoxazole 23 (79.3%). On the other hand, all gram-positive bacterial isolates showed higher sensitivity to nitrofurantoin 27 (93.1%), norfloxacin 24 (82.7%), clindamycin 20 (69%) and ciprofloxacin 19 (65.5%) (Table 5).

**Table 5:** Antimicrobial susceptibility pattern of Gram-positive bacteria (n=29) isolated from urine culture among Asymptomatic female college students at Dessie town, Northeast Ethiopia, January 2021 - March 2021.

| Isolates (n) | Pattern | Antimicrobial agents |  |  |  |  |  |  |  |
| --- | --- | --- | --- | --- | --- | --- | --- | --- | --- |
|  |  | CL | PEN | CAF | CIP | TTC | SXT | F | NOR |
| *S. aureus* (4) | S | 3 (75.0) | 0 (0.0) | 4(100) | 3 (75.0) | 2 (50.0) | 1 (25.0) | 4 (100.0) | 3 (75.0) |
|  | I | 1 (25.0) | 0 (0.0) | 0 (0.0) | 1 (25.0) | 1 (25.0) | 0 (0.0) | 0 (0.0) | 1 (25.0) |
|  | R | 0 (0.0) | 4 (100.0) | 0 (0.0) | 0 (0.0) | 1 (25.0) | 3 (75.0) | 0 (0.0) | 0 (0.0) |
| *S. epidermidis* (1) | S | 1 (100.0) | 0(0.0) | 1(100.0) | 1 (100.0) | 0 (0.0) | 0 (0.0) | 1(100.0) | 1 (100.0) |
|  | I | 0 (0.0) | 0 (0.0) | 0 (0.0) | 0 (0.0) | 0 (0.0) | 1 (100.0) | 0 (0.0) | 0 (0.0) |
|  | R | 0 (0.0) | 1 (100.0) | 0 (0.0) | 0 (0.0) | 1 (100.0) | 0 (0.0) | 0 (0.0) | 0 (0.0) |
| *S. saprophyticus* (24) | S | 16 (66.7) | 1 (4.2) | 9(37.5) | 15 (62.5) | 7 (29.2) | 3 (12.5) | 22 (91.7) | 20 (83.3) |
|  | I | 4 (16.7) | 0 (0.0) | 0 (0.0) | 5 (20.8) | 5 (20.8) | 1 (4.2) | 2 (8.3) | 0 (0.0) |
|  | R | 4 (16.7) | 22 (91.7) | 15 (62.5) | 4 (16.7) | 12 (50.0) | 20 (83.3) | 0 (0.0) | 4 (16.7) |
| Total (29) | S | 20 (69.0) | 1 (3.4) | 14(48.3) | 19 (65.5) | 9 (31.0) | 4 (13.7) | 27 (93.1) | 24 (82.7) |
|  | I | 5 (17.2) | 0 (0.0) | 0 (0.0) | 6 (20.6) | 6 (20.6) | 2 (6.8) | 2 (6.8) | 1 (3.4) |
|  | R | 4 (13.8) | 28 (96.6) | 15(51.7) | 4 (13.7) | 14(48.2) | 23 (79.3) | 0 (0.0) | 4 (13.8) |

CL = Clindamycin, E = Erythromycin, PEN = Penicillin, CAF = Chloramphenicol, CIP = Ciprofloxacin, TTC = Tetracycline, SXT = Trimethoprim-Sulfamethoxazole, F = Nitrofurantoin, NOR = Norfloxacin, R = Resistant, I = Intermediate, S = Sensitive, NT = Not tested

### Multiple drug resistance patterns of the isolates

Overall, all bacterial isolates were resistant to at least two antimicrobial agents whereas 54 (94.7%) isolates were resistant to three or more antimicrobial agents. Multidrug resistance (MDR) was seen in 50 (87.7%) of all isolated bacterial uropathogens. Around 85.7% of Gram negative and 89.6% of gram-positive bacteria showed multidrug resistance for the tested antimicrobial drugs (Table 6).

**Table 6:** Multi drug resistance patterns of bacterial isolates (n=57) from Asymptomatic female college students at Dessie town, Northeast Ethiopia, January 2021 - March 2021.

| Bacterial isolates |  |  |  |  | Antimicrobial resistance pattern |  |  | Total |  |
| --- | --- | --- | --- | --- | --- | --- | --- | --- | --- |
|  |  | R0 | R1 | R2 | R3 | R4 | ≥ R5 |  | MDR |
|  |  | Gram positives |  |  |  |  |  |  |  |
| *S. aureus* |  | 0(0.0%) | 0(0.0%) | 1 (25.0%) | 2 (50.0%) | 1 (25.0%) | 0 (0.0%) | 4 (100.0%) | 3 (75.0%) |
| *S. epidermidis* |  | 0(0.0%) | 0(0.0%) | 0 (0.0%) | 1 (100.0%) | 0 (0.0%) | 0 (0.0%) | 1 (100.0%) | 1 (100.0%) |
| *S. saprophyticus.* |  | 0(0.0%) | 0(0.0%) | 2 (8.3 %) | 2 (8.3 %) | 11 (45.8 %) | 9 (37.5 %) | 24 (100 %) | 22 (91.7%) |
| *Sub total* |  | 0(0.0%) | 0(0.0%) | 3 (10.3 %) | 5 (17.2%) | 12 (41.3) | 9 (31.0) | 29 (100) | 26 (89.6) |
|  |  | Gram negatives |  |  |  |  |  |  |  |
| *E. coli* |  | 0(0.0%) | 0(0.0%) | 0 (0.0%) | 1 (12.5%) | 3 (37.5%) | 4 (50.0%) | 8 (100.0%) | 7 (87.5%) |
| *Citrobacter* |  | 0(0.0%) | 0(0.0%) | 0 (0.0%) | 0 (0.0%) | 1 (50.0%) | 1 (50.0%) | 2 (100.0%) | 2 (100.0%) |
| *Klebsiella ozenae* |  | 0(0.0%) | 0(0.0%) | 0 (0.0%) | 1 (20.0%) | 1 (20.0 %) | 3 (60.0%) | 5 (100.0%) | 4 (80.0%) |
| *Providencia stuarti* |  | 0(0.0%) | 0(0.0%) | 0 (0.0%) | 0 (0.0%) | 0 (0.0%) | 3 (100.0%) | 3 (100.0%) | 3 (100.0%) |
| *Klebsiella rhinoscleromatis* |  | 0(0.0%) | 0(0.0%) | 0 (0.0%) | 0 (0.0%) | 0 (0.0%) | 3 (100.0%) | 3 (100.0%) | 3 (100.0%) |
| *Klebsiella oxytoca* |  | 0(0.0%) | 0(0.0%) | 0 (0.0%) | 0 (0.0%) | 0 (0.0%) | 1 (100.0%) | 1 (100.0%) | 1 (100.0%) |
| *Citrobacter diversus* |  | 0(0.0%) | 1(50.0%) | 0 (0.0%) | 0 (0.0%) | 0 (0.0%) | 1 (50.0%) | 2 (100.0%) | 1 (50.0%) |
| *Acinetobacter spp* |  | 0(0.0%) | 0(0.0%) | 0 (0.0%) | 0 (0.0%) | 0 (0.0%) | 1 (100.0%) | 1 (100.0%) | 1 (100.0%) |
| *Serratia* |  | 0(0.0%) | 0(0.0%) | 0 (0.0%) | 1 (50.0%) | 1 (50.0%) | 0 (0.0%) | 2 (100.0%) | 1 (50.0%) |
| *Klebsiella pneumonia* |  | 0(0.0%) | 0(0.0%) | 0 (0.0%) | 0 (0.0%) | 0 (0.0%) | 1 (100.0%) | 1 (100.0%) | 1 (100.0%) |
| Sub total |  | 0(0.0%) | 0(0.0%) | 0 (0.0%) | 3 (10.7%) | 6 (21.4 %) | 18 (64.2%) | 28 (100.0%) | 24 (85.7) |
| Total |  | 0(0.0%) | 1(1.7%) | 3 (5.2%) | 8 (14.0%) | 18 (31.5%) | 27 (47.3%) | 57 (100.0%) | 50 (87.7%) |

**R2** = Resistance to two, **R3** = Resistance to three, **R4** = Resistance to four, **≥ R5** = Resistance to five and more drugs, **MDR** = multi-drug resistant, based on which MDR definition is applied

## DISCUSSION

The overall prevalence of asymptomatic UTI was 24.6% (95% CI: 18.5-26.1). Our finding is similar with a report in Addis Ababa 23.2% (31). However, higher prevalence were reported from India 40 % (35) and Bangladesh 62.6% (36), which might be due to difference in the study participants. i.e., these studies took a DM patient and hospital attending patients asymptomatic for UTI respectively. Since asymptomatic uropathogens has been maximally associated with DM and other debilitating disease in individuals with low immunity, this can be attributed for high prevalence.

In our study, the overall prevalence of bacterial UTI was 13.5% (95% CI:-10.2-16.6). This is in agreement with studies in Nigeria, 13.8% and 12 % (37, 38). On the other hand, our finding is relatively higher than the findings in Ghana 9.6 % (16). This discrepancy may be due to variation in the number of study participants; i.e., this study took a small number of asymptomatic female students due to high drop-out rate which might decrease the overall prevalence. However, our finding was lower than the reports from Nigeria 46% - 78% (39-41). which might be due to geographical variations. Similarly, higher report of asymptomatic bacteriuria was shown in Ethiopia among diabetic patients (16.7%) (27), HIV patients (18%) (42), pediatric patients (15.9%) (29), which might be attributable to immune suppression of DM and HIV patients which increases susceptibility to bacterial infection. Furthermore, bacteriuria among pediatrics peak during infancy and toilet training period as a result of contamination.

Moreover, the prevalence of asymptomatic Candiduria in this study was 11.1% 95% (CI 8.5-13.7), which is lower than a study done in Nigeria 40.7 (43). Such variation might be attributed to difference in laboratory methods used to identify the microorganism and risk factors with geographical areas.

Unlike most of the study reports in our country and elsewhere in the world, our finding showed majority of the etiological agents for Candiduria were *C. troipcalis (48*.*9)*, followed by *C. albicans (21*.*2)*. Earlier studies report *C. albicans* as the predominant isolate (50-70%) (44). This indicates that there is a paradigm shift toward non-albicans *Candida and* it was further demonstrated that the prevalence of *C. albicans* has significantly declined from 2013-2014 to 2015 (44).

In this study, *S. saprophyticus* (42.1%) followed by *E. coli* (14 %) were the predominant bacteria isolated. *S. saprophyticus causes UTI among* sexually active young women because of displacement from the normal flora of the vagina and perineum into the urethra (32).

This finding is contrary with other studies, which reported *E. coli* as predominant bacterial isolate (27,42,45,40). However, *E. coli* was the most predominant isolate among gram negative bacterial isolates with an isolation rate of 8 (14.0%), which is supported by most of the studies conducted in Hawassa (42) Metu (27) Ghana (16),Nigeria (42.3) (27, 37, 40, 42). The key contributing factor for isolating such a high incidence of E. coli could be presence of E. coli as a faecal flora, which then travels through the genitalia to induce UTI via contamination.; and due to numerous virulence factors used for colonization and invasion of the urinary epithelium such as P-fimbriae or pili adherence factors which mediate the attachment of *E. coli* to uroepithelial cells (32).

Our finding revealed that the frequency of sexual intercourse (>= 3 per week) showed significant association (P<0.001) with the prevalence of UTI. Females having a sexual habit >= 3 per week were 7.907 times more likely to have higher prevalence of asymptomatic UTI than females that have a null sexual frequency habit, which is in agreement with studies in Nigeria (40, 41) This could be due to the fact that female students within this age range (17-30) are sexually active with multi sex partners which may predispose them to UTI (32).

According to our investigation, prevalence of UTI in participants who had genital area washing habit after defecation (P=0.003) and every morning (P=0.008) were also 5.914- and 6.128-times more likely to have higher prevalence of asymptomatic UTI than their counterparts respectively.

This finding agrees with similar reports from Nigeria (40, 46). This could be as a result of poor genital hygienic practices mainly due to low frequency of genital washing and contamination of intestinal flora from feces to the urogenital area.

In our study, 10.5 % of UTI occurred due to bacteria-fungi coinfection. This might be due to the fact that prior cases of urinogenital infection predispose to fungal colonization since the earlier microbial colonizers might have altered the vaginal environment rendering it conducive for infection by Candida species (43).

In the present study, there was no statistically significant association between prevalence of UTI in asymptomatic female college students and age, residence (colleges), student batch, monthly family income, marital status, history of UTI, history of catheterization, history of STDs, and presence of genitourinary abnormalities. Similar finding was reported from Nigeria (40).

Mainly due to the habit of empirical treatment and infrequent bacterial identification and absence of susceptibility testing, antimicrobial resistance among bacterial uropathogens to the commonly used antibiotics become increasing that make clinicians left with very limited choices of drugs for the treatment of urinary tract infection (47). In this study, highest resistance was shown to penicillin (96.6%) and Trimethoprim-Sulfamethoxazole (79.3%) among gram positive bacteria. This could be due to the over use of these drug for many years. On the other hand, lower resistance (higher rate of sensitivity) was observed against nitrofurantoin, norfloxacin, clindamycin and ciprofloxacin. Similar findings have been reported in previous studies done in Dessie (45) and Nigeria (46). The infrequency with which these medications are prescribed could be a possible explanation for such low-level resistance. As a result, they could be used as an alternative to antibiotics in the treatment of UTI.

Amoxicillin-clavulanic acid resistance was quite high among Gram negative bacteria (92.3 %). This could be owing to the widespread availability and indiscriminate use of common medications like amoxicillin-clavulanic acid, which could contribute to an increase in resistance. On the contrary, all tested gram negative isolates showed sensitivity to nitrofurantoin (100.0%), norfloxacin (87.5%) and gentamicin (75.0%) which was also in agreement with the findings of other studies from Hawassa (42) and Nigeria (46) (39).

According to the international standard for definition of drug resistance (48)), multi drug resistance (non-susceptible to ≥ 1 agent in ≥ 3 antimicrobial categories) was observed in 87.7% of the total isolated bacterial uropathogens. This was higher than a study report from Dessie 46.2% (45), Hawassa 78.3% (42) and Addis Ababa 81.1 % (31). This suggests that MDR to routinely used antibiotics in the research location was found to be extremely high. The rise in MDR could be due to the overuse, misuse, and wrong use of antimicrobial drugs in empirical treatment, as well as poor infection control strategy, which could increase the prevalence of resistant microorganisms in the population.

## Conclusions

In this study, a significant prevalence of uropathogens, and an alarmingly high resistance rate of bacterial isolates to commonly used antimicrobial agents were observed among asymptomatic non-pregnant female college students. Nitrofurantoin and norfloxacin were effective for most of gram positive and gram-negative isolates whereas, penicillin, ampicillin, amoxicillin-clavulanic acid and trimethoprim-sulfamethoxazole were less effective for the management of UTI among our study participants. Moreover, significant amount of multi-drug resistance has been shown in most (87.7%) of the bacterial isolates. Frequency of sexual activity and genital area washing habit were significantly associated to have UTI among asymptomatic female college students. Therefore, routine UTI screening, regular health education on the risk of asymptomatic infectious diseases and antimicrobial susceptibility testing should be practiced to avoid the progression of asymptomatic infection into symptomatic UTI.

## Limitations

The study did not include antifungal susceptibility testing due to unavailability of antifungal agent in the market.

## Data Availability

All relevant data are within the manuscript and its Supporting Information files

## List of abbreviations

AHBSc: Alkan health and business science college;
ASB: Asymptomatic bacteriuria;
BMI: Body Mass Index;
CFU: Colony Forming Unit;
CLED: Cystine Lactose Electrolyte-deficient Agar;
CLSI: Clinical and Laboratory Standards Institute;
CNS: Coagulase-negative Staphylococci;
DHSc: Dessie Health Science College;
SDA: Sabouraud dextrose agar;
IUGR: Intrauterine Growth Restriction;
MDR: Multi Drug Resistance;
Mc: Memeherane college;
MIC: Minimum Inhibitory Concentration;
PCR: Polymerase Chain Reaction;
SB: Significant Bacteriuria;
SOPs: Standard Operating Procedures;
SPSS: Statistical Package for the Social Sciences;
TCOM: Tropical college of medicine;
UTI: Urinary Tract Infection;
WHO: World Health Organization

## Acknowledgment

The authors would like to acknowledge Samara University and Wollo University for providing laboratory space and facilities to conduct the experiments. All selected colleges and all study participants are acknowledged for their cooperation during sample collection.

## REFERENCES

1. Haider G, Zehra N, Munir AA, Haider A. Risk factors of urinary tract infection in pregnancy. JPMA The Journal of the Pakistan Medical Association. 2010;60(3):213.

2. Nurullaev R. The role of asymptomatic bacteriuria in epidemiologic study of the urinary tract infection. Likars’ ka sprava. 2004(7):23–5.

3. Ranjan A, Sridhar STK, Matta N, Chokkakula S, Ansari RK. Prevalence of UTI among pregnant women and its complications in newborns. Indian J Pharm Pract. 2017;10:45.

4. August SL, De Rosa MJ. Evaluation of the prevalence of urinary tract infection in rural Panamanian women. PLoS One. 2012;7(10):e47752.

5. Guerra G, de Souza A, da Costa BF, do Nascimento F, Amaral MA, Serafim A. Urine test to diagnose urinary tract infection in high-risk pregnant women. Revista brasileira de ginecologia e obstetricia: revista da Federacao Brasileira das Sociedades de Ginecologia e Obstetricia. 2012;34(11):488.

6. Rosana Y, Ocviyanti D, Karuniawati A, Akhmad SRP. Comparison of microbial pattern causing urinary tract infection in female out-and hospitalized patients in Jakarta. Microbiology Indonesia. 2016;10(1):5-.

7. Iduoriyekemwen N, Sadoh W, Sadoh A. Asymptomatic bacteriuria in HIV positive Nigerian children. Journal of Medicine and Biomedical Research. 2012;11(1):88–94.

8. Bukhary ZA. Candiduria: a review of clinical significance and management. Saudi Journal of Kidney Diseases and Transplantation. 2008;19(3):350.

9. Balachandar M, Pavković P, Metelko Ž. Kidney infections in diabetes mellitus. Diabetologia Croatica. 31(2):85–104.

10. Nayman SA, Özguneş I, Ertem OT, Erben N, Doyuk EK, Tözun M, et al. Evaluation of risk factors in patients with candiduria. Mikrobiyoloji bulteni. 2011;45(2):318–24.

11. Fisher JF. Candida urinary tract infections—epidemiology, pathogenesis, diagnosis, and treatment: executive summary. Clinical infectious diseases. 2011;52(suppl_6):S429–S32.

12. Oluwole OM, Victoria AA. Asymptomatic bacteriuria: occurrence and antibiotic susceptibility profiles among students of a tertiary institution in Ile-Ife, Nigeria. African Journal of Microbiology Research. 2016;10(15):505–10.

13. Mnif MF, Kamoun M, Kacem FH, Bouaziz Z, Charfi N, Mnif F, et al. Complicated urinary tract infections associated with diabetes mellitus: Pathogenesis, diagnosis and management. Indian journal of endocrinology and metabolism. 2013;17(3):442.

14. Moore A, Doull M, Grad R, Groulx S, Pottie K, Tonelli M, et al. Recommendations on screening for asymptomatic bacteriuria in pregnancy. Cmaj. 2018;190(27):E823–E30.

15. Gebremariam G, Legese H, Woldu Y, Araya T, Hagos K, GebreyesusWasihun A. Bacteriological profile, risk factors and antimicrobial susceptibility patterns of symptomatic urinary tract infection among students of Mekelle University, northern Ethiopia. BMC infectious diseases. 2019;19(1):950.

16. Boye A, Dwomoh FP, Morna MT, Dadzie RK. Prevalence of Asymptomatic Bacteriuria in a Sub-population of Tertiary Female Students: Antimicrobial Resistance Patterns and Group-specific Risk Factors. Journal of Medical and Biological Science Research. 2016;2(4):56–64.

17. Nsofor C, Obijuru C, Ozokwor C. Asymptomatic bacteriuria among female students of a tertiary institution in southeast Nigeria. AJRPSB. 2016;4(2):38–44.

18. Diesse JM, Kechia FA, Iwewe YS, Ngueguim AD, Nangwat C, Dzoyem JP. Urinary tract candidiasis in HIV+ patients and sensitivity patterns of recovered Candida species to antifungal drugs in Dschang District Hospital (Cameroon). International Journal of Biological and Chemical Sciences. 2017;11(3):1029–38.

19. Akinjogunla O, Divine-Anthony O, Ajayi A, Etukudo I, Etok I. Asymptomatic Candiduria among Type 1 and 2 Diabetes Mellitus Patients: Risk and Sociodemographic Factors, Prevalence, Virulence Markers and Antifungal Susceptibility. J Pure Appl Microbiol. 2020;14(2):1467–78.

20. Tadesse E, Teshome M, Merid Y, Kibret B, Shimelis T. Asymptomatic urinary tract infection among pregnant women attending the antenatal clinic of Hawassa Referral Hospital, Southern Ethiopia. BMC research notes. 2014;7(1):155.

21. Alemu A, Moges F, Shiferaw Y, Tafess K, Kassu A, Anagaw B, et al. Bacterial profile and drug susceptibility pattern of urinary tract infection in pregnant women at University of Gondar Teaching Hospital, Northwest Ethiopia. BMC research notes. 2012;5(1):197.

22. Derese B, Kedir H, Teklemariam Z, Weldegebreal F, Balakrishnan S. Bacterial profile of urinary tract infection and antimicrobial susceptibility pattern among pregnant women attending at Antenatal Clinic in Dil Chora Referral Hospital, Dire Dawa, Eastern Ethiopia. Therapeutics and clinical risk management. 2016;12:251.

23. Demilie T, Beyene G, Melaku S, Tsegaye W. Urinary bacterial profile and antibiotic susceptibility pattern among pregnant women in North West Ethiopia. Ethiopian journal of health sciences. 2012;22(2).

24. Ali IE, Gebrecherkos T, Gizachew M, Menberu MA. Asymptomatic bacteriuria and antimicrobial susceptibility pattern of the isolates among pregnant women attending Dessie referral hospital, Northeast Ethiopia: A hospital-based cross-sectional study. Turkish journal of urology. 2018;44(3):251.

25. Assefa A, Asrat D, Woldeamanuel Y, Abdella A, Melesse T. Bacterial profile and drug susceptibility pattern of urinary tract infection in pregnant women at Tikur Anbessa Specialized Hospital Addis Ababa, Ethiopia. Ethiopian medical journal. 2008;46(3):227–35.

26. Tsega A, Mekonnen F. Prevalence, risk factors and antifungal susceptibility pattern of Candida species among pregnant women at Debre Markos Referral Hospital, Northwest Ethiopia. BMC Pregnancy and Childbirth. 2019;19(1):1–8.

27. Gutema T, Weldegebreal F, Marami D, Teklemariam Z. Prevalence, antimicrobial susceptibility pattern, and associated factors of urinary tract infections among adult diabetic patients at Metu Karl Heinz Referral Hospital, Southwest Ethiopia. International Journal of Microbiology. 2018;2018.

28. Marami D, Balakrishnan S, Seyoum B. Prevalence, Antimicrobial Susceptibility Pattern of Bacterial Isolates, and Associated Factors of Urinary Tract Infections among HIV-Positive Patients at Hiwot Fana Specialized University Hospital, Eastern Ethiopia. Canadian Journal of Infectious Diseases and Medical Microbiology. 2019;2019.

29. Merga Duffa Y, Terfa Kitila K, Mamuye Gebretsadik D, Bitew A. Prevalence and antimicrobial susceptibility of bacterial uropathogens isolated from pediatric patients at yekatit 12 hospital medical college, Addis Ababa, Ethiopia. International journal of microbiology. 2018;2018.

30. Debalke S, Cheneke W, Tassew H, Awol M. Urinary tract infection among antiretroviral therapy users and nonusers in Jimma University Specialized Hospital, Jimma, Ethiopia. International journal of microbiology. 2014;2014.

31. Woldemariam HK, Geleta DA, Tulu KD, Aber NA, Legese MH, Fenta GM, et al. Common uropathogens and their antibiotic susceptibility pattern among diabetic patients. BMC infectious diseases. 2019;19(1):43.

32. Cheesbrough M. District laboratory practice in tropical countries, part 2: Cambridge university press; 2006.

33. Borman AM, Fraser M, Johnson EM. CHROMagarTM Candida Plus: A novel chromogenic agar that permits the rapid identification of Candida auris. Medical Mycology. 2021;59(3):253–8.

34. Institute CaLS. CLSI.Performance Standards for Antimicrobial susseptablity Testing. ClSI Supplement M100. USA: Wayne, PA; 2020.

35. Nongrum S, Thaledi S, Singh VA, Narang V, Mehta S, Garg R, et al. Association of uropathogens with asymptomatic urinary tract infection in diabetes mellitus patients. Int J Curr Microbiol App Sci. 2016;5(10):355–61.

36. Hossain MJ, Siddiqi A, Rahman MM, Khan KN, Imtiaj A. Prevalence of Urinary Tract Infection of Female Patients in Northern Bangladesh. 2017.

37. Chijioke A. Nsofor., et al. ”Asymptomatic Bacteriuria Among Female Students of a Tertiary Institution in Southeast Nigeria”. EC Bacteriology and Virology Research. 2016;2:106–12.

38. Frank-Peterside N, Wokoma E. Prevalence of asymptomatic bacteriuria in students of University of Port Harcourt Demonstration Secondary School. Journal of Applied Sciences and Environmental Management. 2009;13(2).

39. Alao FO, Akintunde FA. Asymptomatic Bacteriuria among Students of Bellstech, Ota, Nigeria. Pacific J Sci Technol. 2012;13:342–7.

40. Mbata C, Aleru C, Azike C, Adewoye M, Emeka E, Ugwueze V. Incidence of asymptomatic bacteriuria amongst female students residing in school hostels of Rivers state University of Science and Technology, Port Harcourt. Int J Curr Res Med Sci. 2016;2(10):55–9.

41. Christiana AO, Perpetua ON, Obasi OS. Burden of Urinary Tract Infection (UTI) among Female Students: South Eastern Nigeria Side of the Story. International Journal of TROPICAL DISEASE & Health. 2015:1-7.

42. Tessema NN, Ali MM, Zenebe MH. Bacterial associated urinary tract infection, risk factors, and drug susceptibility profile among adult people living with HIV at Haswassa University Comprehensive Specialized Hospital, Hawassa, Southern Esthiopia. Scientific Reports. 2020;10(1):1–9.

43. Enweani I, Ogbonna C, Kozak W. The incidence of candidiasis amongst the asymptomatic female students of the University of Jos, Nigeria. Mycopathologia. 1987;99(3):135–41.

44. Rathor N, Khillan V, Sarin S. Nosocomial candiduria in chronic liver disease patients at a hepatobilliary center. Indian Journal of Critical Care Medicine: Peer-reviewed, Official Publication of Indian Society of Critical Care Medicine. 2014;18(4):234.

45. Alemu M, Belete MA, Gebreselassie S, Belay A, Gebretsadik D. Bacterial Profiles and Their Associated Factors of Urinary Tract Infection and Detection of Extended Spectrum Beta-Lactamase Producing Gram-Negative Uropathogens Among Patients with Diabetes Mellitus at Dessie Referral Hospital, Northeastern Ethiopia. Diabetes, Metabolic Syndrome and Obesity: Targets and Therapy. 2020;13:2935.

46. Christiana AO, ONP, Obasi aOS. Burden of Urinary Tract Infection (UTI) among Female Students. International Journal of TROPICAL DISEASE & Health 2016;12(2):1–7.

47. Beyene G, Tsegaye W. Bacterial uropathogens in urinary tract infection and antibiotic susceptibility pattern in jimma university specialized hospital, southwest ethiopia. Ethiopian journal of health sciences. 2011;21(2):141–6.

48. Magiorakos A-P, Srinivasan A, Carey Rt, Carmeli Y, Falagas Mt, Giske Ct, et al. Multidrug-resistant, extensively drug-resistant and pandrug-resistant bacteria: an international expert proposal for interim standard definitions for acquired resistance. Clinical microbiology and infection. 2012;18(3):268–81.

